# Impact of CancelRx on Discontinuation of Controlled Substance Prescriptions

**DOI:** 10.1101/2021.01.12.21249700

**Authors:** Taylor L. Watterson, Jamie A Stone, Aaron Gilson, Roger Brown, Ka Z Xiong, Anthony Schiefelbein, Edmond Ramly, Peter Kleinschmidt, Michael Semanik, Lauren Craddock, Samantha Pitts, Taylor Woodroof, Michelle Chui

**Affiliations:** University of Wisconsin-Madison School of Pharmacy, Madison, WI, USA; University of Wisconsin-Madison School of Nursing, Madison, WI, USA; Wisconsin Department of Health Services, Madison, WI, USA; University of Wisconsin School of Medicine and Public Health, Madison, WI, USA; University of Wisconsin-Madison College of Engineering, Madison, WI, USA; Compass Dermatopathology, Inc., San Diego, CA, USA; Johns Hopkins Medicine, Baltimore, MD, USA

## Abstract

**Objective:** To assess how controlled substance medication discontinuations were communicated over time

**Data Sources:** Secondary data from a midwestern academic health system electronic health record and pharmacy platform were collected 12-months prior to CancelRx implementation and for 12-months post implementation.

**Study Design:** The study utilized an interrupted time series analysis (ITSA) to capture the proportion of controlled substance medications that were cancelled in the clinic’s electronic health record and also cancelled in the pharmacy’s dispensing software. The ITSA plotted the proportion of successful cancellation messages over time, particularly after the health system’s implementation of CancelRx, a novel technology.

**Data Collection/Extraction:** Data were extracted from the EHR and pharmacy records for patients aged 18+ who had a controlled substance discontinued by a health system provider. Information collected included patient demographics, drug information (name, dose), and dates discontinued in the clinic and pharmacy records.

**Principal Findings:** After CancelRx implementation there was a significant increase in the proportion of discontinued controlled substance medications that were communicated to the pharmacy.

**Conclusions:** This study demonstrates the role that technology can play in promoting controlled substance policy and medication safety.

## INTRODUCTION

In March 2018, the National Institute on Drug Abuse reported that more than 120 people die in the United States every day due to opioid overdoses.^1^ The Centers for Disease Control and Prevention (CDC) cites this as a “serious national crisis” and estimates that the total economic burden of prescription opioid misuse amounts to $78.5 billion in the United States each year (including the cost of healthcare, lost productivity, addiction treatment, and criminal justice system involvement).^2^ More recent data from the CDC’s National Vital Statistics Report in December 2018 stated that the number of annual drug overdose deaths has increased 54% between 2011 and 2016 (increasing from 41,340 to 63,632 deaths per year).^3^ The CDC reported that the top 10 drugs involved in these overdose deaths belonged to three drug classes:

- Pain Medications (opioids): fentanyl, hydrocodone, methadone, morphine, and oxycodone
- Benzodiazepines: alprazolam and diazepam
- Stimulants: cocaine and methamphetamine derivatives

In light of this crisis, agencies including state governments are promoting and requiring the use of electronic prescribing (e-prescribing) of all controlled prescriptions to reduce dispensing errors as well as drug diversion and misuse.^4,5^ When a medication is prescribed for a patient, a hand-written prescription creates opportunities for errors, such as incorrect transcription from the clinic electronic health record (EHR) to the prescription or incorrect translation from the prescription to the pharmacy system or vial label.^4^ Additionally, a hand-written controlled substance prescription provides opportunities for individuals to tamper with or alter the contents from the prescriber’s original intent. Such alterations can include changing the strength, dose, quantity, or directions for use.^6^ Electronic prescribing of controlled substances streamlines and controls all aspects of the drug dispensing process – sending the prescription directly from the clinic EHR to the pharmacy dispensing software and minimizing the opportunities for unintentional error or intentional abuse.

However, e-prescribing also creates vulnerabilities in the dispensing of controlled substances. For example, federal law prohibits the inclusion of refills on prescriptions for Schedule II controlled substances (those deemed as having a currently accepted medical use but a high potential for abuse). he need for new prescriptions for these continuous orders may yield the same medication/strength/dose being listed several times on a patient’s medication list (both in the clinic EHR and pharmacy dispensing software). These duplicate orders may be confusing to decipher or add unnecessary noise throughout the prescribing and dispensing process. These vulnerabilities are further exacerbated when the dose of the medication is tapered or changed, resulting in confusion about which order is the most recent among a long list of the same product. These changes, as well as a general lack of integration, often lead to medication list discrepancies between clinic EHR and pharmacy dispensing platforms.^7–9^

In addition to vulnerabilities in the ordering and dispensing processes, e-prescribing of controlled substances (EPCS) also presents unanticipated opportunities for drug diversion, misuse, and potential abuse. A common anecdote describes a scenario in which a patient is seen at the clinic and is prescribed a controlled substance. Knowing that the prescriber will transmit a prescription electronically to their desired pharmacy, the patient has a colleague stationed at the pharmacy to retrieve the medication as soon as it is filled. However, after the prescription is sent to the first pharmacy, the patient exclaims that they have changed their mind and requests that the order, instead be sent to another pharmacy (often a different chain or network than the first). After the second prescription is sent, they either have another colleague waiting there, or go to retrieve the medication themselves. These patients often pay cash or utilize coupons to avoid insurance billing errors or flags. Additionally, by filling the prescription the same day, these patients avoid Prescription Drug Monitoring Program (PDMP) flags or warnings at the pharmacy (pharmacy reporting to the PDMP is reported every 24 hours).^10,11^ In these scenarios, even if the prescriber delegates a clinic staff member to contact the first pharmacy to cancel the original prescription, it may be too late when the patient’s representative is already there to retrieve the medication as soon as it is filled.

These vulnerabilities emphasize the need for efficient and effective communication of medication cancellations between prescriber clinics and community pharmacies. There are various ways in which these medication cancellations can be communicated, including notes attached to new e-prescriptions, telephone calls from clinic staff, faxed messages, or novel health information technology (health IT) such as CancelRx. CancelRx is a health IT functionality that utilizes the same electronic pathway as e-prescriptions.^12–19^ A third party vendor (SureScripts) sends a communication directly from the clinic EHR to the pharmacy dispensing software, but instead of authorizing the fill of a medication, it details its cancellation. Depending on the community pharmacy integration, a CancelRx can even identify and halt the processing of a previously e-prescribed prescription, working behind the scenes to ensure a prescription is effectively cancelled. In a pilot study by Pitts et. al. CancelRx demonstrated a 92.4% successful rate in which a medication was discontinued in the prescriber’s EHR and successfully discontinued in the pharmacy’s dispensing software.^19^ Similarly, a study by Chui et. al, showcased not only a 93% success rate for CancelRx communication medication discontinuations, but also a drastic reduction in the amount of time required for a medication to be discontinued in the pharmacy software after it was cancelled at the clinic.^20^

CancelRx has an added benefit, not just for patient safety and minimizing medication list discrepancies but thwarting the potential dispensing of prescription medications used for non-medical purposes. This presents an opportunity to assess the impact of CancelRx on the cancellation and dispensing of prescription drugs that may be contributing to the opioid crisis. By electronically communicating medications that are discontinued in the physician’s office, this health IT functionality can potentially minimize the number of extraneous or unnecessary controlled medications dispensed and available to patients as well as reduce potential for diversion. This analysis is timely and relevant given CancelRx’s inclusion in CMS Meaningful Use Criteria (to be implemented by 2020) as well as ongoing need to address the serious opioid crisis.^12,14^

### Objectives

The main objective of the study was to measure the impact of CancelRx on reducing controlled substance medication list discrepancies between the clinic EHR and the pharmacy management software, specifically opioids, benzodiazepines, and stimulants. This was assessed by evaluating the percentage of controlled substance prescriptions successfully discontinued in the EHR and pharmacy management software over time. A secondary study objective was to compare the impact of CancelRx on reducing medication list discrepancies for controlled substances and non-controlled substances. This was done by assessing the percentages of controlled substance and non-controlled substance prescriptions successfully discontinued in the EHR and pharmacy management software over time. Finally, a third objective was to assess the impact of CancelRx on the length of time (in days) between medication discontinuation in the clinic EHR and discontinuation in pharmacy dispensing software.

## METHODS

This study capitalized on the opportunity for a natural experiment, when an academic health system, UW Health, in Wisconsin implemented CancelRx in October of 2017. Prior to CancelRx, when a prescriber decided to discontinue a medication, they would document the change in the clinic EHR. The prescriber could potentially send a message to the pharmacy documenting the medication change (call, fax, or e-prescription note), or delegate the task to an appropriate member of the clinic staff (often through EHR inbox messaging). The clinic staff could then follow through with communicating the cancellation message to the pharmacy. Once a cancellation message was received at the pharmacy, the pharmacy staff was responsible for identifying, cancelling, and documenting the appropriate prescription. This process provided numerous opportunities for human or technology vulnerabilities that yielded the cancellation message incomplete.

With CancelRx, when a prescriber decides to discontinue a medication, he or she still documents the change in the EHR. With this new health IT functionality, however. The prescriber can relay the cancellation information directly to the pharmacy via a CancelRx – importantly, a prescriber can still opt out of sending a cancellation message to the pharmacy, but the default/nonresponse setting is that a CancelRx is sent. A third-party vendor, SureScripts, attempts to send the cancellation message from the clinic EHR to the community pharmacy dispensing system. This process is enhanced when the discontinued medication was previously e-prescribed; SureScripts can follow the same electronic signature or automatically identify the prescription in the pharmacy’s dispensing software and halt it from being further dispensed or processed. In the event that SureScripts is unable to send the cancellation message electronically, it still routes a message to the appropriate clinic staff to follow-up with the necessary pharmacy manually. This instantaneous process allows for clinic staff to be informed of issues throughout the discontinuation process. Additionally, even if a prescription cannot be matched automatically within the pharmacy’s dispensing system, a member of the pharmacy staff can still manually match the cancellation message to the appropriate prescription to ensure that it is discontinued within the system. This type of notification also alerts the pharmacy staff to changes in a patient’s medication list in addition to the immediate behind-the-scenes removal of the prescription.

UW Health underwent a phased approach of e-prescribing Schedule II medications implementation. Data extracted from the EHR included information regarding the patient (only those 18+), clinic encounter, discontinued drug, and dates of discontinuation. Because the health system in the study implemented mandatory e-prescribing of controlled substances, the research team was able to capitalize on the SureScript e-prescription electronic identification to match medications that were discontinued in the clinic and supposed to be discontinued in the pharmacy both before and after CancelRx implementation. In addition to the use of the e-prescription signature, orders were also matched on patient gender, drug description (name, strength), ordering department, and a drug discontinuation time at the pharmacy within 72-hours of when it was discontinued at the clinic.

The research team extracted data from the health system EHR (Epic) regarding controlled substance medications discontinued in the outpatient clinics at 12-months prior to CancelRx implementation and for 12-months post implementation (allowing for a 4-week a priori burn-in period).

An interrupted time series analysis (ITSA) was used to determine the impact of CancelRx on controlled substance medication list discrepancies over time. Specifically, the ITSA allowed the research team to compare the proportion of controlled substances that were discontinued clinic EHR and successfully discontinued in the pharmacy dispensing software within 72-hours. The study utilized Prais-Winsten estimation, meant to address the serial correlation of type AR(1) in a linear model, with analysis conducted in STATA.^21–24^ A one-week period constituted a measurement in the time series.

Furthermore, time-to-discontinuation event analyses were also conducted to compare the length of time between EHR and pharmacy system discontinuation before and after CancelRx implementation. The time to discontinuation was aggregated and averaged for each week in the study period and compared over time using Regression with Newey-West standard errors.^25,26^

This study was approved by the University of Wisconsin-Madison Institutional Review Board.

## RESULTS

### Outcome 1: Percentage of Successful Controlled Substance Discontinuations Over Time

Figure 1 illustrates the ISTA for all controlled substances, where the x-axis depicts time (in weeks) with a vertical line at Week 56 indicating CancelRx implementation. The y-axis depicts the proportion of medications that were successfully discontinued in both the clinic and pharmacy systems. The pre-intervention period showed a moderate trend increase (0.474% per week) prior to implementation of CancelRx. After CancelRx implementation, there was an immediate and significant increase in the proportion of successful medication discontinuations, an increase of 77.7% (adjusted for orders, p = 0.00). The post-implementation slope remained very stable at 0.03%.

**Figure 1.**
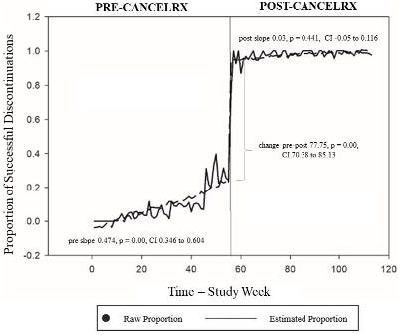
Successful Controlled Substance Discontinuations Pre- and Post-CancelRx Implementation

### Outcome 2: Percentage of Successful Discontinuations Over Time for Controlled Substances and Non-Controlled Substances

Figure 2 illustrates the ISTA comparing the proportion of successful medication discontinuations for controlled substances compared to non-controlled substances. The ISTA illustrated that, while the trend for non-controlled substances remained fairly stable in the year prior to CancelRx implementation (pre-implementation slope 0.02%), the proportion of successful controlled substance discontinuations gradually increased until almost converging at approximately 30% success.

**Figure 2.**
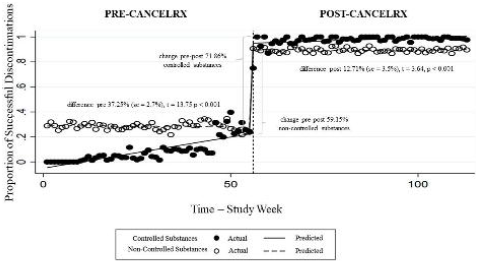
Successful Discontinuations Pre- and Post-CancelRx Implementation, Controlled Substance vs Non-Controlled Substance

Immediately following CancelRx implementation, the proportion of successfully discontinued medications significantly increased for both controlled and non-controlled substances. However, the percentage increase was greater for controlled substances than non-controlled substances, and this difference persisted throughout the year following CancelRx implementation. The trends for both controlled substances and non-controlled substances were stable in the post-CancelRx implementation period.

### Outcome 3: Time to Discontinuation Between Clinic and Pharmacy Over Time

The third outcome of the study compared the average amount of time between when a medication was discontinued in the clinic EHR and when it was discontinued in the pharmacy dispensing software.

Given that the clinic EHR data contained both date and time stamps for when a medication was discontinued, whereas the pharmacy data contained only the date, it was necessary to address this inconsistency. Therefore, when matching and combining the clinic and pharmacy datasets, prescriptions that were discontinued on the same day (in both the clinic and pharmacy) were coded as 0 days, while discontinuations in the pharmacy the following day were coded as 1 day, etc. When these orders were aggregated for an entire week, the results were often represented as a fraction of days. To make the results easier to comprehend, they are presented in Figure 3 as hours (i.e., 0.5 days = 12 hours) and represent an average across cases.

**Figure 3.**
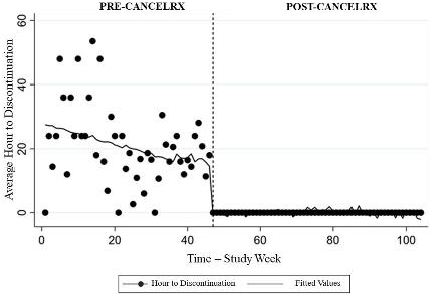
Average Time to Controlled Substance Discontinuations Pre- and Post-CancelRx Implementation

**Figure 4.**
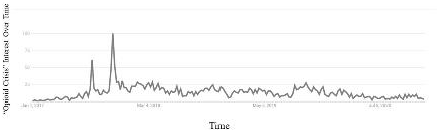
“Opioid Epidemic” Interest Over time Data Source: Google Trends Numbers represent search interest relative to the highest point on the chart for the given region and time. A value of 100 is the peak popularity for the term.

In the year prior to CancelRx implementation, the time required for a medication to be discontinued in the pharmacy after it had been cancelled at the clinic varied. Using regression with Newey-West standard errors, the time to discontinuation was 27.43 hours (se = 4.48, t=6.12, p = 0.00, CI 18.53 - 36.31). Prior to CancelRx implementation, there were several weeks in which all orders were discontinued the same day (represented with y-values = 0) or weeks in which the average time was over 2 days (y-values > 48). The predicted trend-line seemed to gradually decrease in the weeks prior to CancelRx implementation.

After CancelRx implementation, medication discontinuations were all completed on the same day (all values were = 0) with a very stable trend and little variation.

## DISCUSSION

Overall, implementation of CancelRx showed a marked improvement in the percentage of medications that were successfully discontinued in the pharmacy after being discontinued at the clinic. The addition of this novel technology effectively minimized the incidence of medication list discrepancies at the pharmacy. However, the study results must be situated in the environmental context of October 2017 when CancelRx was implemented.

### Pre-CancelRx

The pre-intervention period demonstrated a gradual increase in the number of medications that were successfully discontinued in both the clinics and pharmacies leading up to CancelRx implementation. There were several organization-wide factors that may have contributed to this upward trend. First, in the year prior to CancelRx, [Affiliation] began incrementally implementing the electronic transmission of controlled substance prescriptions to pharmacies (specifically EPCS). Although prescriptions had long before been “electronically” documented in the clinic’s EHR, prior to EPCS prescribers would print-out and hand the patients a paper copy of their prescription or mail prescriptions to the pharmacy to be dispensed. Prior to EPCS, clinic staff may have faced difficulty tracking exactly where a patient filled their prescription. This could lead to confusion regarding who clinic staff should notify when medications were stopped or discontinued. Even if clinic staff members attempted to contact the patient’s preferred pharmacy (as documented in the EHR), the patient could have filled the prescription elsewhere and the pharmacy staff would have no record of the medication being discontinued. This could create distrust or frustration for clinic staff who attempted to reduce patient medication list discrepancies by contacting the preferred pharmacy and met only with additional questions.

As the health system implemented EPCS in the year prior to CancelRx, clinic staff could ensure where prescriptions were being filled (based on where they were electronically sent) and, therefore, may have been more confident that when they attempted to contact the pharmacies, they would have the medication on the patent’s record. Additionally, the EPCS implementation facilitated a growing culture and discussion surrounding Schedule II controlled substances that may have contributed to increased staff awareness and ensuring the need to communicate when medications were stopped or discontinued. For example, when prescriptions can be electronically transmitted to the pharmacy via the patient’s EHR, there is potentially more importance on having a concise, clean, and up-to-date medication list without duplicate or outdated prescriptions. This “clean” list would ensure that the most recent and correct prescriptions are renewed. An organizational culture emphasizing the importance of up-to-date medication lists may lead clinic staff to increasingly contact the pharmacy regarding outdated or previously discontinued prescriptions.

Beyond the scope of the singular [Affiliation] organization, Wisconsin enacted legislation that required prescribers and clinic staff to check the Prescription Drug Monitoring Program (PDMP) prior to writing a prescription for a Controlled Substance.^27^ Occurring in April 2017, this statute emphasized the use of the statewide registry that documents the patient, drug name, drug strength, quantity and pharmacy information, as well as third-party payment methods used when opioids, benzodiazepines, and stimulants are filled. Mandated use of this tool may have heightened awareness surrounding the prescribing and discontinuation of controlled substances and led to the increasing trend in the months preceding CancelRx implementation.

The timeline leading up to CancelRx implementation also coincides with events on a national scale. On October 26, 2017, President Trump declared the opioid crisis a public health emergency. This public declaration led to an increase in public awareness surrounding opioid and controlled substance prescribing, as evident by the Google Trend line shown in the Figure below (indicating the number of times the term “opioid crisis” was searched, or interest over time). Once again, this announcement likely increased the general conversation surrounding controlled substance prescriptions, and motivated clinics, prescribers, and pharmacies alike to have accurate and up-to-date medication lists for patients using these medications.

### Post-CancelRx

After CancelRx implementation, there was a significant increase in the communication of medication cancellation messages. Although the influence of local, state, and national environment surrounding controlled substances is still important to consider, the drastic and sustained success also seems attributable to the CancelRx functionality itself. CancelRx was designed to work “behind the scenes” and automate the previously manual process, eliminating the need for clinic staff to intervene, except in cases in which the prescription was not electronically transmitted to the pharmacy (such as prescriptions written prior to EPCS), the CancelRx could not be sent (such as the pharmacy had not turned on the functionality) or the pharmacy was unable to find a match for the discontinued prescription (such as the prescription had been transferred out to another pharmacy). This demonstrated that not only was the health IT functionality able to take over tasks and reduce workload for clinic staff and MAs, but that they were able to perform the task more efficiently.

CancelRx clearly impacted the way that clinics and pharmacies communicated regarding the discontinuation of controlled substance medications. However; CancelRx may have also improved coordination and communication within clinics. Implementation of the CancelRx functionality reduced barriers to communicating cancellation messages and may have increased prescriber and clinic awareness and motivation to address this issue. The ability to quickly and reliably discontinue medications from a patient’s medication list in the EHR may have prompted prescribers and clinic staff to “clean up” patient’s profiles, which ensured that only most recent and up to date medications were listed on the patient’s records. This helps to facilitate a common organizational mental model around the way that controlled substance medications should be documented, especially when documenting medication discontinuations.

An example of ways in which CancelRx may have improved communication and reduced barriers in the clinic is when prescribers are requested to “cover” their colleagues and prescribe continuing doses of controlled substances. If a patient or patient’s pharmacy requests a refill requested of a controlled substance, [Affiliation] policy indicated that the clinic staff should first verify that the patient is eligible for a refill by checking the PDMP and noting the last fill date of the prescription. If the patient is due for a refill, the clinic staff sends the request off to the patient’s provider or original prescriber. If the provider is not in the clinic, the request may be forwarded to another prescriber in the clinic who is “covering” for the intended recipient. This process exposes the patient to potential vulnerabilities.

The first potential vulnerability is if the patient’s medication list in the EHR still has a medication on the patient’s profile that was actually discontinued. The clinic staff and prescriber may look at the patient’s chart and not know or see that the prescriber actually wanted the patient to stop taking the medication. Unknowingly, the covering prescriber could continue the medication therapy by authorizing refills. In this sense, CancelRx and the emphasis on ensuring an up-to-date and accurate medication lists, reduces the unintended supply of controlled substances reaching patients.

A second potential vulnerability is when the patient’s care plan includes tapering the doses down of controlled substance medications. The clinic’s covering provider may not know or be able to find documentation of the plan in the patient’s EHR to slowly reduce the patient’s dose or directions and simply continue the patient’s previous therapy. CancelRx, and the clinic’s mental model surrounding discontinued medications facilitates safe patient handoffs across providers.

### Impact of CancelRx on Controlled Substances and Non-Controlled Substances

CancelRx functionality was implemented to address medication cancellation messages for all prescriptions, and not just for controlled substances. Therefore the research team assessed the differences between communication of medication discontinuations for controlled substances and non-controlled substances.

In comparing the results of the controlled substances and non-controlled substances post-CancelRx the overall levels for the controlled substances were actually higher than that of the non-controlled substances. This may be in part to the organizational, state, and national emphasis placed on controlled substance prescribing (and discontinuation). This perhaps confirms the notions of a shared mental model and organizational and cultural awareness surrounding this issue and the desire to minimize unintentional supply of controlled substances reaching patients. To put another way, prescribers and clinic staff were motivated to ensure up-to-date medication lists in the post-CancelRx period not only because of the priority and attention given to the topic but also because the functionality reduced barriers to communication.

## Limitations

A study must be considered in light of its limitations. This study has limited generalizability because it occurred within a single health system with both outpatient clinics and affiliated pharmacies. Additionally, these community pharmacies had access to a patient’s EHR, to further investigate questions regarding medication discontinuations or patient’s care plans. Future studies should assess the impact of CancelRx for patients who fill prescriptions at community pharmacies outside of a singular health system to determine the impact on medication list discrepancies and communication between prescribers and pharmacists.

A second limitation is the confounding effects of the environment in which CancelRx implementation took place. Situated amongst ECPS roll outs, PDMP statues, and attention at the national level, the effects of CancelRx may be inflated. However, the results are still convincing given the notable and sustained increase in medication discontinuations communicated after CancelRx implementation.

## Conclusions

This study illustrated the impact of a novel technology, CancelRx, on communicating medication discontinuations between clinics and pharmacies. Namely, this study assessed the changes in medication list discrepancies when the prescriptions being discontinued were controlled substances. In light of the opioid epidemic, it is crucial for both clinics and pharmacies to have up-to-date medication lists, not only for the safety of patients but also to prevent abuse and misuse within their communities. This study demonstrates the role that technology can play in promoting controlled substance medication safety.

## Data Availability

The data underlying this article cannot be shared publicly for the privacy of individuals that participated in the study; the dataset contains protected health information. The data underlying this article were provided by UW Health by permission. Data will be shared on reasonable request to the corresponding author with permission of UW Health.

